# PREVALENCE OF ABDOMINAL OBESITY AND ITS ASSOCIATION WITH CARDIOVASCULAR RISK AMONG THE ADULT POPULATION IN BURKINA FASO

**DOI:** 10.1101/2021.01.25.21250457

**Authors:** Kadari Cisse, Sékou Samadoulougou, Mady Ouedraogo, Seni Kouanda, et Fati Kirakoya-Samadoulougou

**Author notes:** Corresponding author: Kadari Cissé, Institut de Recherche en Sciences de la Santé (IRSS), Ouagadougou, 03 BP 7192, Burkina Faso.

## Abstract

**Objective:** The objective of this study is to determine the prevalence of abdominal obesity and its associated factors in Burkina Faso. We hypothesize that there is a high burden of abdominal obesity and it is significantly associated with sociodemographic and cardiovascular risk factors.

**Design:** We performed secondary analysis of the survey conducted in Burkina Faso using the World Health Organization (WHO) STEPwise approach.

**Setting:** The study was conducted in Burkina Faso with all 13 regions of the country included.

**Participants:** Our study involved 4308 adults of both sexes aged 25 to 64 years.

**Main outcome:** Our primary outcome was the abdominal obesity which was could defined using a cut-off point of waist circumference (WC) of ≥94 cm for men and ≥80 cm for women.

**Results:** The overall age-standardized prevalence of abdominal obesity was 22.5% (95% CI: 21.3–23.7). This age-standardized prevalence was 35.9% (95% CI: 33.9–37.9) among women and 5.2% (95% CI: 4.3–6.2) among men (p < 0.001). In urban areas, the age-standardized prevalence of abdominal obesity was 42.8% (95% CI: 39.9–45.7) and 17.0% (95%CI: 15.7–18.2) in rural areas (p < 0.001). The overall age-standardized prevalence of very high WC (WC ≥102 cm for men and ≥88 cm for women) was 10.2% (95%CI: 9.3–11.1). According to the National Institute for Health and Clinical Excellence (NICE) BMI–WC matrix, which combines the body mass index (BMI) and WC to define different levels of cardiovascular health risk, 14.6% of adult Burkinabè had an increased cardiovascular health risk.

**Conclusion:** Our study shows a high prevalence of abdominal obesity among the adult population in Burkina Faso. These findings suggest that the measurement of WC should be systematically incorporated in Burkina Faso primary healthcare centers for the early detection of high cardiovascular risk in order to reduce levels of premature death.

**STRENGTHS AND LIMITATIONS OF THIS STUDY:**

➢ This is the first national representative study on abdominal obesity in the context of an emerging epidemiological transition in Burkina Faso.
➢ A recommended cut-off point was used to define abdominal obesity among the adult population in Burkina Faso, which we found to be associated with “intermediate” cardiovascular risk factors.
➢ The waist circumference and risk factors used in this study were measured using the standard approach proposed by the WHO [1]. However, some risk factors such as physical inactivity, alcohol consumption, and type of fat were self-reported and may therefore be affected by information bias.
➢ This study was a cross-sectional study and must not be considered to make causal inference.

**Target journal:** https://bmjopen.bmj.com/

## INTRODUCTION

Obesity is an increasing public health issue worldwide, particularly in developing countries. Globally, 603.7 million adults were found to be obese in 2015, and this number has been rising since 1980 [2]. Obesity is most often assessed using the body mass index (BMI) [3,4], which is recognized to increase in parallel with the risk of cardiovascular diseases such as coronary heart disease or stroke [5]. However, the BMI provides limited information on body fat distribution, which is related to metabolic risk [5,6]. The majority of fat is stored in subcutaneous adipose tissue; however, in some individuals, excessive amounts may be accumulated intra-abdominally (visceral fat) [3]. The visceral accumulation of body fat is due to genetic factors [7–13], neuroendocrine perturbations [14], and environment and lifestyle factors [15]. The combination of the overconsumption of energy-dense food and a sedentary lifestyle is well known to play a role in the accumulation of visceral fat [3,16,17]. The BMI alone seems insufficient to assess the distribution of body fat and evaluate the cardiometabolic risk among adults with excess of adiposity because the BMI fails to capture the cardiometabolic risk related to abdominal obesity [18]. Abdominal obesity is responsible for increased risk of insulin resistance, type 2 diabetes mellitus, metabolic syndrome, cardiovascular diseases, cancers, chronic respiratory diseases, and all-cause mortality [3,19–23].

Three anthropometric proxies are commonly used to assess abdominal obesity: the waist circumference (WC), waist-to-height ratio, and waist-to-hip ratio. Most studies on abdominal obesity have used WC as the defining criterion [3,24,25]. The WC is known to be sensitive to visceral fat accumulation [26]. Individuals with a larger WC have more abdominal fat than those with a smaller WC [26]. The threshold of WC used to define abdominal obesity depends on ethnic group and world regions. For sub-Saharan Africa (SSA), the World Health Organization (WHO) defined abdominal obesity by fixing sex specific WC cut-off points at ≥94 cm for men and ≥80 cm for women.[27] This cut-off point appears to increase the risk of metabolic complications. Furthermore, the risk of cardiometabolic disorders is “substantially increased” when the WC is ≥102 cm among men and ≥88 cm among women [27]. Using the WHO definition, Wong et *al*.[28] reported that nearly half (41.5%) of the global adult population was abdominally obese and that this prevalence is rising worldwide, including in low- and middle-income countries in SSA. However, there is a paucity of data on abdominal obesity in SSA as previously highlighted by Wong et al. [27]. Some studies have been conducted in South Africa [29], Kenya [30], Uganda [31], Nigeria [32], and Cote d’Ivoire [33]. Except for that of Kabwama et *al*. [28] in Uganda, most of these studies were performed in local areas and, therefore, do not provide country-level estimates of the prevalence of abdominal obesity. The new consensus of the International Atherosclerosis Society (IAS) and the International Chair on Cardiometabolic Risk (ICCR) Working Group on Visceral Obesity, published in March 2020, recognized that decreasing WC is a critical target to reduce the adverse health risks for both sexes [18].

A recent population-based nationwide study in Burkina Faso showed a high proportion (one out of five) of adults had abnormal weight [34]. Some local studies on abdominal obesity have previously been conducted. Thus, in Northern Ouagadougou, in 2010, Zeba et *al*. [35] noted that the prevalence of abdominal obesity was 12.5%. In their study of specific populations in Bobo-Dioulasso, Yameogo and *al*. [36] found that 64.9% of diabetics were abdominally obese. These studies do not provide information on the nationwide prevalence of abdominal obesity. As is the case in many other developing countries, the management of cardiometabolic risk factors is becoming a public health challenge in Burkina Faso [37]. The added value of our study comes from our focus on the prevalence of abdominal obesity and its associated risk factors among the adult population in Burkina Faso using data from the first national non-communicable disease risk factors survey.

## METHODS

### Study design, setting and population

We performed a secondary analysis of data from a cross-sectional survey, the national WHO STEPS survey, which aim of assessing the risk factors for NCDs, and was carried out between 26 September and 18 November 2013 in Burkina Faso. Burkina Faso is located in the SSA region in West Africa, covering a surface area of 272,960 km2 with 20,870,060 habitants in 2019 with life expectancy at birth of 61.8 years. The proportion of the population living in urban areas increased (according to the results of last national population census) from 12.7% in 1985 to 22.7% in 2006. The epidemiological profile is dominated by infectious diseases, with an increasing burden of non-communicable diseases (NCDs), including cardiovascular diseases, resulting in the country facing a double burden of disease and a progressive change of the pattern of diseases. The data used in our study were collected from a representative sample of adults between 25 and 64 years old. The study was designed to provide estimates at the national, regional, and place of residence (urban/rural) levels. Participants were selected using a three-stage cluster stratified sampling method. A total of 240 enumeration areas (EAs) that corresponded to the cluster were sampled. In each EA, 20 households were randomly selected. One respondent was identified in each households studied [38].

### Data collection

Data were collected electronically on a Personal Digital Assistant (PDA) and consisted of face-to-face interviews, which were conducted after obtaining informed consent from the respondent. Risk factors linked to NCDs, such as physical activities and biochemical parameters, of the subjects selected to participate in the survey were collected using a STEPS instrument. The survey procedures consisted of 3 steps: the first step focused on sociodemographic information, behavioral measures, and questions on physical activity, food hygiene, oral health, and knowledge of NCD risk factors. Behavioral measures related to the consumption of tobacco and alcohol were also collected. The second step measured the following physical parameters: the height was measured using a portable measuring rod for participants without shoes and without a hat. Weight was measured using an electronic weighing scale (SECA®) with the person being weighed lightly clothed and without shoes. The WC (umbilical perimeter) was measured using a measuring tape applied directly to the skin along the axillary line, midway between the lower base of the last rib and the iliac crest of each side; the measurement was taken only once and reported to the nearest 0.1 cm. Participants were ineligible for waist measurement if pregnant. Blood pressure was measured using an electronic sphygmomanometer. The third step was to measure blood sugar level and blood cholesterol from a capillary blood sample using an electronic device, CardioChek.

### Variables of interest

#### Outcome variable

In our study, the dependent variable was abdominal obesity, which was measured using the WC for each of the study participants. For the primary outcome, i.e., the cut-off, a WC of ≥94 cm for men and ≥80 cm for women was used. According to the WHO, this cut-off is associated with an increased risk of cardiometabolic complications. To estimate the proportion of adults who need to reduce their weight, we used a cut-off of WC ≥102 cm for men and ≥88 cm for women as a secondary outcome, as recommended by Lean et *al*. [39] This corresponds to a very high WC [40]. At this cut-off, the risk of developing cardiometabolic complications increases substantially.[5] Thus, the WC collected during the STEPS survey was categorized into a binary variable: 0 “without high WC <94 cm (men) and <80 cm (women)”, 1 “with high WC ≥94 cm (men) and ≥80 cm (women)” for the primary outcome. A similar categorization was used for the secondary outcome: 0 “without high WC <102 cm (men) and <88 cm (women)”, 1 “with high WC ≥102 cm (men) and ≥88 cm (women)”.

#### Explanatory variables

To take into consideration the influence of potential predictors on the WC, we selected three classes of associated factors based on previous studies [29,31,41]. Among these risk factors, there were sociodemographic, behavioral, and metabolic factors:

✓ **Sociodemographic factors:** Regarding the demographic factors, we categorized all of the variables. Among these variables are the following: age (25–34, 35–44, 45–54, 55–64 years), sex (male/female), highest education level (no level, primary, secondary or higher), marital status (unmarried/married), professional status (wage earner, self-employed, unemployed) and place of residence (urban/rural).
✓ **Behavioral factors:** The behavioral variables that could influence our dependent variable and were selected for our study are the following: smoking status (yes/no), alcohol use (yes/no), number of fruits or vegetables eaten per day, type of fat intake and physical activities (high, moderate, and low intensity).
✓ **Metabolic factor:** We used the height and weight of the individuals to obtain their BMI and then categorized it into four groups (BMI < 18.5 = underweight; 18.5 ≤ BMI < 25 = normal; BMI ≥25 = overweight; BMI ≥ 30 = obesity). We also checked the association between abdominal obesity and other cardiovascular intermediate risk factors such as high blood pressure (HBP) as defined by the WHO (systolic blood pressure ≥ 140 mmHg or diastolic blood pressure ≥ 90 mmHg) and diabetes (capillary fasting blood glucose above or equal 6.1 mmol/L) and hypercholesterolemia (capillary total cholesterol above or equal 5.2 mmol/L).

### Cardiovascular health risk assessment

To assess cardiovascular health risk, we used the National Institute for Health and Clinical Excellence (NICE) BMI–WC matrix approach, which combines the BMI and WC values to define different levels of health risk [40]. This matrix has been used in South Africa to determine the prevalence of adults with an increased health risk [29]. The new consensus of the International Atherosclerosis Society (IAS) and the International Chair on Cardiometabolic Risk (ICCR) Working Group on Visceral Obesity recommended the combination of BMI and WC to identify people with high health risk [18]. We also added high blood pressure, diabetes, and hypercholesterolemia as independent variables (adjusted for sociodemographic and behavioral characteristics of the study population) to check their association with abdominal obesity in this study.

### Statistical method

We first described the characteristics of the study population through a weighted analysis to take the sampling design into account. The standardized prevalence of abdominal obesity was presented using the sociodemographic characteristics of the study population and cardiovascular risk factors. Standardization was achieved using the age structure of the adult population of Burkina Faso in 2013 [42]. We implemented a modified Poisson regression model using a generalized estimating equation to derive prevalence ratios (PRs) while taking into account clustering of observations. We performed univariate and multivariable analyses of individual-level covariates. The PRs were calculated with 95% confidence intervals. Statistical significance was accepted at the 5% level (p < 0.05).

## RESULTS

### Sociodemographic characteristics of the study population

A total of 4308 participants with valid WC and BMI data were included in this study. The mean age was 38.5 ± 11.2 years and 41% of participants were between 25 and 34 years old, 52.4% were female, and 74.1% were living in rural areas. The people who had not attended formal school represented 78.6% of the participants and 87.2% were married (see Table 1 for more details).

**Table 1:** Background characteristics of the study population.

| Characteristics | Number | Percentage* |
| --- | --- | --- |
| **Sex** | 4308 |  |
| Women | 2151 | 52.4 |
| Men | 2157 | 47.6 |
| **Age group, years** | 4308 |  |
| 25–34 | 1917 | 41.0 |
| 35–44 | 1078 | 27.8 |
| 45–54 | 796 | 19.2 |
| 55–64 | 517 | 12.0 |
| **Marital status** | 4303 |  |
| Single | 601 | 12.8 |
| Married | 3702 | 87.2 |
| **Highest completed level of education** | 4300 |  |
| No formal school | 3380 | 78.6 |
| Primary school | 652 | 14.9 |
| Secondary or higher | 268 | 6.5 |
| **Profession** | 4303 |  |
| Wage earner | 210 | 5.2 |
| Self-employed | 3088 | 68.9 |
| Unemployed | 1005 | 25.9 |
| **Residence** | 4308 |  |
| Urban | 878 | 25.9 |
| Rural | 3430 | 74.1 |
| **Risk factors** |  |  |
| **Current smoker** | 4307 |  |
| No | 3760 | 88.4 |
| Yes | 547 | 11.6 |
| **Current drinker** | 4307 |  |
| No | 3107 | 72.1 |
| Yes | 1200 | 27.9 |
| **Fruit and vegetable intake** | 4010 |  |
| <5 | 3819 | 95.7 |
| ≥5 | 191 | 4.3 |
| **Type of fat most commonly used** | 4197 |  |
| Vegetable oil | 2620 | 63.3 |
| Butter, lard or fat, margarine | 1157 | 27.2 |
| None or other | 420 | 9.5 |
| **Physical activity** | 4307 |  |
| Intense | 2639 | 60.6 |
| Moderate | 1110 | 25.5 |
| Low | 558 | 13.9 |
| BMI class | 4308 |  |
| Underweight | 494 | 11.9 |
| Normal | 3095 | 70.2 |
| Overweight | 548 | 13.4 |
| Obese | 171 | 4.5 |
| HBP |  |  |
| No | 3441 | 76.9 |
| Yes | 864 | 20.1 |
| Diabetes |  |  |
| No | 4052 | 95.4 |
| Yes | 193 | 4.6 |
| Hypercholesterolemia |  |  |
| No | 4190 | 97.8 |
| Yes | 93 | 2.2 |

\*weighted percentage

### Prevalence of abdominal obesity

Out of 4308 participants, 876 had abdominal obesity. As shown in Table 2, the overall age-standardized prevalence of abdominal obesity (primary outcome) was 22.5% (95% CI: 21.3–23.7). This age-standardized prevalence was 35.9% (95% CI: 33.9–37.9) among women and 5.2% (95%CI: 4.3–6.2) among men (p < 0.001). The prevalence increase with age group was as follows: 19.6% (95% CI: 17.1–22.4) for 25–34 years, 24.0% (95% CI: 20.8–27.6) for 35–44 years, 25.7% (95% CI: 21.5–30.3) for 45–54 years; 25.3% (95% CI: 20.8–30.6) for 55–64 years. No difference was observed regarding marital status, and the prevalence of abdominal obesity among married people was 22.5% (95% CI: 21.2–23.8) and 23.6% (95% CI: 19.7–27.5) among unmarried people. The prevalence of abdominal obesity was 19.4% (95% CI: 18.1–20.7) among people who had not attended formal school and 33.4% (95% CI: 29.5–37.2) for people who attended primary school and 45.8% (95% CI: 39.8–51.8) for people who attended at least secondary school. In urban areas, the age-standardized prevalence of abdominal obesity was 42.8% (95% CI: 39.9–45.7) and 17.0% (95% CI: 15.7–18.2) in rural areas (p < 0.001). Among women, the prevalence of abdominal obesity in urban areas was 64.4% (95CI: 60.1–68.7) versus 27.8% (95% CI: 25.6–29.9) in rural areas, and among men, it was 14.8% (95% CI: 11.3–18.4] in urban areas and 3.1% (95% CI: 2.3–3.9) in rural areas.

**Table 2:** Age-standardized prevalence of abdominal obesity according to characteristics of the study population.

| Characteristics | Men |  | Women |  | Total |  |
| --- | --- | --- | --- | --- | --- | --- |
|  | n | Prev [95%CI] | n | Prev [95%CI] | n | Prev [95%CI] |
| **All participants** | **2157** | **5.2 [4.3–6.2]** | **2151** | **35.9 [33.92–37.9]** | **4308** | **22.5 [21.3–23.7]** |
| **Sex** |  |  |  |  |  |  |
| Women |  |  |  |  | 2151 | 35.9 [33.9–37.9] |
| Men |  |  |  |  | 2157 | 5.2 [4.3–6.2] |
| **Age group, years** |  |  |  |  |  |  |
| 25–34 | 915 | 3.5 [2.3–5.4] | 1002 | 32.4 [28.2–37.0] | 1917 | 19.6 [17.1–22.4] |
| 35–44 | 539 | 8.8 [6.1–12.5] | 539 | 38.3 [32.9–44.0] | 1078 | 24.0 [20.8–27.6] |
| 45–54 | 407 | 6.6 [4.4–9.8] | 389 | 44.4 [37.2–51.9] | 796 | 25.7 [21.5–30.3] |
| 55–64 | 296 | 9.7 [6.2–14.8] | 221 | 43.4 [35.9–51.2] | 517 | 25.3 [20.8–30.6] |
| **Marital status** |  |  |  |  |  |  |
| Single | 326 | 4.6 [2.1–7.1] | 275 | 38.2 [31.6–44.9] | 601 | 23.6 [19.7–27.5] |
| Married | 1828 | 5.1 [4.1–6.2] | 1874 | 36.1 [33.8–38.3] | 3702 | 22.5 [21.2–23.8] |
| **Highest completed level of education** |  |  |  |  |  |  |
| No formal school | 1595 | 3.8 [2.8–4.7] | 1785 | 31.6 [29.5–33.7] | 3380 | 19.4 [18.1–20.7] |
| Primary school | 389 | 6.1 [3.6–8.7] | 263 | 54.5 [47.9–61.1] | 652 | 33.4 [29.5–37.2] |
| Secondary or more | 165 | 21.9 [15.7–28.3] | 103 | 63.9 [54.4–73.4] | 268 | 45.8 [39.8–51.8] |
| **Profession** |  |  |  |  |  |  |
| Wage earner | 152 | 17.8 [11.4–24.2] | 58 | 64.3 [54.1–74.5] | 210 | 44.2 [37.8–50.7] |
| Self-employed | 1914 | 4.1 [3.2–4.9] | 1174 | 30.8 [28.2–33.5] | 3088 | 19.1 [17.6–20.6] |
| Unemployed | 90 | 9.1 [1.8–16.5] | 915 | 39.8 [36.6–42.9] | 1005 | 26.4 [22.7–30.1] |
| **Residence** |  |  |  |  |  |  |
| Urban | 407 | 14.8 [11.3–18.4] | 471 | 64.4 [60.1–68.7] | 878 | 42.8 [40.0–45.7] |
| Rural | 1750 | 3.1 [2.3–3.9] | 1680 | 27.8 [25.6–29.9] | 3430 | 17.0 [15.7–18.2] |
| **Risk factors** |  |  |  |  |  |  |
| **Current smoker** |  |  |  |  |  |  |
| No | 1612 | 5.5 [4.4–6.6] |  |  | 3760 | 22.6 [21.0–23.8] |
| Yes | 545 | 4.2 [2.4–6.0] |  |  | 547 | 9.6 [0.0–20.5] |
| **Current drinker** |  |  |  |  |  |  |
| No | 1494 | 5.0 [3.9–6.1] | 1613 | 36.9 [34.5–39.3] | 3107 | 22.9 [21.5–24.3] |
| Yes | 663 | 6.0 [4.1–7.8] | 537 | 34.3 [30.3–38.4] | 1200 | 21.9 [19.5–24.4] |
| **Fruit and vegetable intake** |  |  |  |  |  |  |
| <5 | 1909 | 5.0 [4.0–6.0] | 1910 | 35.7 [33.6–37.9] | 3819 | 22.3 [21.0–23.6] |
| ≥5 | 83 | 4.6 [0.2–9.0] | 108 | 43.5 [34.6–52.5] | 191 | 26.5 [21.1–31.9] |
| Type of fat most commonly used |  |  |  |  |  |  |
| Vegetable oil | 1313 | 7.2 [5.8–8.6] | 1307 | 40.8 [38.1–43.5] | 2620 | 26.1 [24.5–27.7] |
| Butter, lard or fat, margarine | 534 | 1.7 [0.7–2.8] | 623 | 27.9 [24.4–31.4] | 1157 | 16.4 [14.4–18.5] |
| None or other | 232 | 2.1 [0.3–4.0] | 188 | 30.3 [23.7–36.8] | 420 | 17.9 [14.1–21.6] |
| Physical activity |  |  |  |  |  |  |
| Intense | 1437 | 4.2 [3.2–5.2] | 1202 | 36.2 [33.4–38.9] | 2639 | 22.2 [20.6–23.8] |
| Moderate | 468 | 4.3 [2.5–6.1] | 642 | 32.6 [28.9–36.2] | 1110 | 20.2 [18.0–22.4] |
| Low | 252 | 12.4 [8.3–16.5] | 306 | 41.3 [35.8–46.9] | 558 | 28.7 [25.1–32.3] |
| BMI class |  |  |  |  |  |  |
| Underweight | 189 | 1.4 [0.0–3.4] | 305 | 8.4 [5.4–11.4] | 494 | 5.3 [3.4–7.2] |
| Normal | 1652 | 1.5 [0.9–2.0] | 1443 | 27.1 [24.8–29.4] | 3095 | 15.8 [14.5–17.2] |
| Overweight | 265 | 19.8 [15.1–24.5] | 283 | 85.9 [81.9–89.8] | 548 | 57.1 [54.0–60.2] |
| Obese | 51 | 63.6 [51.1–76.1] | 120 | 95.9 [92.4–99.4] | 171 | 82.0 [76.2–87.7] |
| HBP |  |  |  |  |  |  |
| No | 1686 | 4.0 [3.0–4.9] | 1755 | 33.1 [30.8–35.3] | 3441 | 20.4 [19.0–21.7] |
| Yes | 470 | 9.3 [6.6–11.9] | 394 | 47.7 [42.4–52.9] | 864 | 30.9 [27.7–34.1] |
| Diabetes |  |  |  |  |  |  |
| No | 2023 | 5.1 [4.1–6.1] | 2029 | 35.5 [33.4–37.6] | 4052 | 22.2 [20.9–23.4] |
| Yes | 101 | 6.4 [2.1–10.7] | 92 | 44.4 [34.4–54.4] | 193 | 27.9 [21.9–33.9] |
| Hypercholesterolemia |  |  |  |  |  |  |
| No | 2112 | 5.0 [4.1–6.0] | 2078 | 34.5 [32.5–36.6] | 4190 | 21.6 [20.4–22.8] |
| Yes | 33 | 17.6 [5.2–30.0] | 60 | 78.7 [68.2–89.2] | 93 | 52.2 [44.1–60.3] |

HBP: high blood pressure; BMI: body mass index, Prev: age-standardized prevalence; n = number of adults; CI: confidence interval

Regarding the secondary outcome, it is known that a WC of ≥102 for men and ≥88 for women (very high WC) substantially increases the risk of cardiovascular disease, and according to the expert consensus of the WHO, people at and above this cut-off point should follow weight-loss strategies to reduce this risk. At this cut-off point, the age-standardized prevalence of very high WC was 10.2% (95% CI: 9.3–11.1). It was 16.9% (95% CI: 15.3–18.5) among women and 1.6% (95% CI: 1.1–2.1%) among men. Regarding residence, we found that the age-standardized prevalence of very high WC was 25.3% (95% CI: 22.7–28.0) in urban areas and 6.0% (95% CI: 5.2–6.8) in rural areas (p < 0.001).

### Factors associated with abdominal obesity

Table 3 shows the results of multivariable analysis. The main variables found to be associated with abdominal obesity are sex, age group, marital status, education, profession, and residence. When the analysis was stratified by sex, we found that among women, age group, marital status, education, and residence were significantly associated with abdominal obesity, while among men, only age group and residence were significantly associated with abdominal obesity (see Table 3 for more details).

**Table 3:** The GEE analysis results for abdominal obesity.

| Characteristics | Male |  | Female |  | Total |  |
| --- | --- | --- | --- | --- | --- | --- |
|  | cPR [95%CI] | aPR [95%CI] | cPR [95%CI] | aPR [95%CI] | cPR [95%CI] | aPR [95%CI] |
| **Sex** |  |  |  |  |  |  |
| Women |  |  |  |  | 1 | 1 |
| Men |  |  |  |  | 0.15 [0.12–0.18] | 0.15 [0.11–0.19] |
| **Age group. years** |  |  |  |  |  |  |
| 25–34 | 1 | 1 | 1 | 1 | 1 | 1 |
| 35–44 | 1.73 [1.08–2.78] | 2.14 [1.25–3.68] | 1.29 [1.13–1.48] | 1.25 [1.09–1.44] | 1.33 [1.15–1.54] | 1.30 [1.13–1.49] |
| 45–54 | 1.69 [0.97–2.95] | 1.54 [0.78–3.07] | 1.53 [1.32–1.77] | 1.54 [1.31–1.80] | 1.49 [1.26–1.76] | 1.51 [1.28–1.78] |
| 55–64 | 2.09 [1.27–3.44] | 1.82 [0.97–3.41] | 1.57 [1.32–1.85] | 1.62 [1.33–1.98] | 1.39 [1.15–1.68] | 1.64 [1.34–2.00] |
| **Marital status** |  |  |  |  |  |  |
| Single | 1 | 1 | 1 | 1 | 1 | 1 |
| Married | 1.50 [0.85–2.65] | 1.46 [0.86–2.50] | 1.04 [0.88–1.24] | 1.17 [1.00–1.38] | 1.27 [1.02–1.39] | 1.21 [1.03–1.43] |
| **Highest completed level of education** |  |  |  |  |  |  |
| No formal school | 1 | 1 | 1 | 1 | 1 | 1 |
| Primary school | 1.36 [0.80–2.28] | 1.19 [0.64–2.21] | 1.29 [1.10–1.50] | 1.26 [1.07–1.48] | 1.00 [0.84–1.18] | 1.21 [1.02–1.42] |
| Secondary or more | 3.39 [2.06–5.64] | 1.51 [0.73–3.13] | 1.24 [0.95–1.62] | 1.09 [0.83–1.44] | 1.02 [0.77–1.35] | 1.18 [0.92–1.52] |
| **Profession** |  |  |  |  |  |  |
| Wage earner | 1 | 1 | 1 | 1 | 1 | 1 |
| Self-employed | 0.30 [0.19–0.47] | 0.80 [0.40–1.60] | 0.65 [0.49–0.84] | 0.77 [0.58–1.02] | 0.70 [0.48–1.00] | 0.71 [0.55–0.92] |
| Unemployed | 0.65 [0.34–1.22] | 1.16 [0.61–2.22] | 0.73 [0.57–0.94] | 0.82 [0.62–1.09] | 1.69 [1.20–2.37] | 0.80 [0.63–1.03] |
| **Residence** |  |  |  |  |  |  |
| Urban | 1 | 1 | 1 | 1 | 1 | 1 |
| Rural | 0.22 [0.15–0.33] | 0.31 [0.20–0.49] | 0.42 [0.36–0.50] | 0.48 [0.40–0.58] | 0.36 [0.31–0.43] | 0.50 [0.42–0.60] |
| **Risk factors** |  |  |  |  |  |  |
| **Current smoker** |  |  |  |  |  |  |
| No | 1 | 1 | 1 | 1 | 1 | 1 |
| Yes | 0.71 [0.46–1.11] | 0.91 [0.55–1.54] | 1.41 [0.87–2.29] | 2.87 [2.37–3.47] | 0.17 [0.11–0.27] | 0.77 [0.46–1.29] |
| **Current drinker** |  |  |  |  |  |  |
| No | 1 | 1 | 1 | 1 | 1 | 1 |
| Yes | 1.21 [0.83–1.77] | 0.89 [0.59–1.34] | 1.01 [0.87–1.16] | 0.94 [0.82–1.08] | 0.86 [0.74–1.00] | 0.97 [0.84–1.11] |
| **Fruit and vegetable intake** |  |  |  |  |  |  |
| <5 | 1 | 1 | 1 | 1 | 1 | 1 |
| ≥5 | 1.05 [0.43–2.60] | 1.26 [0.45–3.49] | 1.18 [0.95–1.47] | 1.24 [0.98–1.56] | 1.23 [0.92–1.64] | 1.27 [0.98–1.63] |
| Type of fat most commonly used |  |  |  |  |  |  |
| Vegetable oil | 1 | 1 | 1 | 1 | 1 | 1 |
| Butter, lard or fat, margarine | 0.29 [0.15–0.57] | 0.37 [0.17–0.83] | 0.93 [0.81–1.06] | 0.96 [0.84–1.11] | 0.91 [0.77–1.06] | 0.88 [0.76–1.02] |
| None or other | 0.34 [0.14–0.80] | 0.54 [0.22–1.34] | 0.93 [0.72–1.20] | 0.92 [0.68–1.24] | 0.78 [0.57–1.06] | 0.82 [0.59–1.14] |
| Physical activity |  |  |  |  |  |  |
| Intense | 1 | 1 | 1 | 1 | 1 | 1 |
| Moderate | 1.01 [0.62–1.64] | 0.88 [0.54–1.45] | 0.92 [0.80–1.07] | 0.90 [0.78–1.03] | 1.19 [1.01–1.41] | 0.89 [0.77–1.03] |
| Low | 2.83 [1.87–4.28] | 1.70 [1.06–2.71] | 1.07 [0.91–1.26] | 0.94 [0.80–1.10] | 1.51 [1.27–1.80] | 1.03 [0.88–1.20] |
| HBP |  |  |  |  |  |  |
| No | 1 | 1 | 1 | 1 | 1 | 1 |
| Yes | 2.45 [1.69–3.54] | 1.84 [1.20–2.83] | 1.37 [1.22–1.54] | 1.20 [1.06–1.36] | 1.41 [1.23–1.62] | 1.30 [1.14–1.47] |
| Diabetes |  |  |  |  |  |  |
| No | 1 | 1 | 1 | 1 | 1 | 1 |
| Yes | 1.54 [0.85–2.78] | 1.42 [0.83–2.41] | 1.16 [0.92–1.46] | 1.07 [0.84–1.38] | 1.16 [0.90–1.50] | 1.17 [0.92–1.49] |
| Hypercholesterolemia |  |  |  |  |  |  |
| No | 1 | 1 | 1 | 1 | 1 | 1 |
| Yes | 3.45 [1.46–8.13] | 1.66 [0.76–3.61] | 1.77 [1.41–2.21] | 1.50 [1.19–1.91] | 2.39 [1.82–3.13] | 1.52 [1.18–1.94] |

GEE: general estimation equation; cPR: crude prevalence ratio; aPR: adjusted prevalence ratio

### Abdominal obesity and cardiovascular health risk

Using the NICE BMI–WC matrix to assess health risk, we found that the prevalence of “increased risk”, “high risk”, and “very high risk” to health was 6.8%, 4.6%, and 3.2%, respectively. The prevalence of at least increased risk to health was 14.6% (see Table 4 for more details) and 83.6% of study population was classified as having “no health risk”. After adjusting for the sociodemographic and behavioral characteristics of study population, we found that abdominal obesity was significantly associated with HBP and hypercholesterolemia. Indeed, as shown in Table 3, the prevalence of abdominal obesity was 70% (aPR: 1.30; 95% CI: 1.14–1.47) higher among people with HBP compared to those without. This association was also reported among both women and men when the multivariable analysis by sex was stratified. Thus, the prevalence of abdominal obesity was 61% higher for women and 37% higher for men with HBP compared to those without (see Table 3 for more details). Furthermore, the prevalence of abdominal obesity was 52% (aPR: 1.52; 95%CI: 1.18–1.94) higher among participants with hypercholesterolemia. This association was persistent even if we adjusted the model with BMI.

**Table 4:** Cardiovascular health risk assessment using the NICE BMI–WC matrix.

|  |  | Waist circumference (WC) |  |  |
| --- | --- | --- | --- | --- |
|  |  | Low | High | Very high |
| Body Mass Index (BMI) class | Underweight (<18.5 kg/m<sup>2</sup>) | 11.1 | 0.5 | 0.2 |
|  | Normal (18.5 to <25 kg/m<sup>2</sup>) | 59.9 | 8.0 | 2.4 |
|  | Overweight (25 to <30 kg/m<sup>2</sup>) | 5.8 | 3.9 | 3.8 |
|  | Obese (30 to <40 kg/m<sup>2</sup>) | 0.5 | 0.8 | 2.7 |
|  | Morbid obese (40 kg/m<sup>2</sup> or more) | 0.1 | 0.0 | 0.4 |

*Low = WC of <80 cm for women and <94 cm for men; High = WC of ≥80 and <88 cm for women and ≥94 and <102 cm for men; Very high = WC of ≥88 for women and ≥102 cm for men*
No increased risk Increased risk High risk Very high risk
*The prevalence of increased risk, high risk, and very high risk of health was 6.8%, 4.6% and 3.2%, respectively. The prevalence of at least increased risk was 14.6%.*

## DISCUSSION

In this study, the prevalence of abdominal obesity was estimated at 22.5%. The prevalence of abdominal obesity was significantly higher among individuals who were older, female, had primary school as their highest level of education, and living in urban areas. Abdominal obesity was also associated cardiovascular risk factors such as hypertension and hypercholesterolemia. Using a cut-off point for WC of ≥102 cm for men and ≥88 cm for women, which is considered to substantially increase cardiovascular risk, we found that nearly one in ten (10.2%) adults had very high WC. Based on the NICE matrix, nearly one in six adults in Burkina Faso had an increased risk of cardiovascular diseases.

In the literature, different cut-off points for WC have been used to evaluate abdominal obesity. Indeed, using a cut-off point for WC of ≥94 cm for men and ≥80 cm for women, a recent meta-analysis by Wong et *al*. [28] reported that the global prevalence of abdominal obesity was 41.5% [95% CI: 39.9–43.2%]. This prevalence is higher than those reported in our study using the same WC cut-off point. It is known from Wong et *al*. [28] that the prevalence of abdominal obesity is higher among populations of high-income countries and Caucasians [28]. In the context of SSA, different prevalences of abdominal obesity have been reported depending on the cut-off point used and study population. Indeed, Awolabi et *al*. [29] noted that the prevalence of abdominal obesity in South Africa was 67% when using the same cut-off as Wong et *al*. [28]. The prevalence of abdominal obesity in Kenya was found to be 52.0% [30]. A study in Nigeria showed a prevalence of 30.1% [32]. Yayehd et *al*. [43] noted that the prevalence of abdominal obesity was 48.8% in semi-urban areas in Togo. Using the cut-off point for WC of ≥102 cm for men and ≥88 cm for women, Kabwama et *al*. [31] showed that the prevalence of abdominal obesity in Uganda was 11.8%, which is similar to those reported in our study using the same cut-off point. In SSA, abdominal obesity is seen as a sign of wealth, affluence, respect, and dignity [31]. This harmful perception of abdominal obesity among the adult population of SSA presents a challenge for the implementation of weight management strategies. Considering the mediating role of abdominal obesity in the development of diabetes and cardiovascular and other chronic diseases combined with the high prevalence and harmful perception of abdominal obesity in many SSA countries, there is an urgent need for strategies to raise awareness regarding the health implications of abdominal obesity [29]. Its high prevalence among some populations in SSA, including in Burkina Faso, presage a future epidemic of cardiometabolic complications if no effective action is taken. The current levels of abdominal obesity among the adult population in Burkina Faso should invoke policy makers to draft effective weight management strategies in the high-risk groups reported in this study.

In this study, we report a significantly higher prevalence of abdominal obesity among individuals who are older, women, have a higher education level, and are urban residents. As shown by Wong et *al*. [28], the global prevalence trends of abdominal obesity show it is significantly higher among older individuals, female subjects, urban residents, Caucasians, and populations of higher income level countries. Kabwama et *al*. [31] also found that the prevalence of abdominal obesity was significantly higher among women, individuals who attended secondary school, and urban residents in Uganda. The higher prevalence of abdominal obesity reported among women in many studies in both high- and low-income countries appears to have numerous causes [28,31,44]. Firstly, differences in the deposition of abdominal fat for men and women is well known to be influenced by differences in the level of steroid hormones between the sexes—these hormones drive body structure during adolescence. Secondly, as shown by some researchers, the deposition of body fat in adolescents appears to be influenced by genetic and environmental factors, which results in women having increased susceptibility to fat accumulation compared to men [28,31,44]. Thirdly, the amount of abdominal fat tends to increase in women since it is affected by each of the pregnancy phases and the post-menopausal distribution of body fat. The high prevalence of abdominal obesity in older individuals compared to younger is, on one hand, due to the fact that older individuals are typically physically inactive and expend less energy than young adults. On the other hand, this situation might also be explained by the well-known lower basal metabolic among older adults which, at this age, contributes to the accumulation of excess body fat due to the increased ratio of energy intake to expenditure. However, the onset of abdominal obesity at an early age appears to be associated with an increased risk of mortality [45]. This suggests the necessity for NCD policies to prevent abdominal obesity among the young [28]. In our study, education level and professional status were significantly associated with abdominal obesity. The education level is a proxy indicator of higher socioeconomic status, which is known to increase the risk of abdominal obesity [46,47]. This relation appears to disappear when adjusted for physical inactivity [46], suggesting the need to promote a healthy lifestyle among adults with a high level of education. We reported a high prevalence of abdominal obesity among urban residents. Socioeconomic status and urbanization were associated with dietary habit, which corresponds to excess fat and calorie intake. They is also known to be associated with low physical activities and stressful conditions, which tend to increase cortisol secretion and abdominal obesity risk.

In this study, we reported a high proportion (14.6%) of the adult population to be at high risk of body fat-related cardiometabolic complications in Burkina Faso. Using the same approach (NICE BMI–WC), Owolabi et *al*. [29] reported that nearly 67% of their study participants in South Africa had at least an increased risk to health. South Africa is at an advanced stage of epidemiological transition with a heavy burden of overweight and abdominal obesity [29]. Regarding our findings, it seems that Burkina Faso is at early stage of the growing trend of overnutrition, which is one of the characteristics of the epidemiological transition. Such a situation offers a unique opportunity to control the prevalence of abdominal obesity and its related health risk before they reach those in middle- and high-income countries. This might be achieved through voluntary weight management strategies, which are needed for at least one in five (22.7%) adults in Burkina Faso (if considering the WHO expert consensus). It might also be achieved through routine measurements of WC combined with BMI in clinical practice, as has been recommended by the IAS and ICCR Working Group on Visceral Obesity [18]. WC is known to be associated with cardiovascular and all-cause mortality regardless of adjustment for BMI [48,49]. WC is a simple anthropometric measurement which can be easily conducted in settings with limited resources and might help in screening for cardiometabolic risk [18]. In our study, we found that abdominal obesity was significantly associated with hypertension and hypercholesterolemia but not with diabetes. The same finding had been noted by Owolabi et *al*. [29]. In addition, abdominal obesity is known to be an important mediator of insulin resistance and endothelial dysfunction [50]. Our results might be explained by the fact that many abdominally obese people may not yet have the biological symptoms of insulin resistance. The association between hypercholesterolemia and abdominal obesity appeared to be conserved even in the model adjusted for BMI categories. This finding suggests that the waist is an indicator of the total cholesterol level among adults. This result also calls for the use of WC measurements in primary healthcare since it is a “vital sign” in clinical practice [18] and total cholesterol is difficult to evaluate in limited-resource settings at the primary care level.

## CONCLUSION

Our study showed a high prevalence of abdominal obesity among adults in Burkina Faso and a high proportion of adults with abdominal obesity who require weight management strategies to prevent the occurrence of cardiometabolic complications. The prevalence was significantly higher among women, the elderly, have a higher education level and adults who resided in urban areas. This finding could invoke policy makers to improve weight-loss strategies in the country. Promoting the adoption of a healthy lifestyle and dietary habits might curb the rising household-level and health-system-related costs of cardiometabolic complications associated with abdominal obesity. This is crucial in limited-resource settings, such as in Burkina Faso, to prevent premature health deterioration and encourage sustainable economic growth in a country currently at an early stage of epidemiological and demographic transition.

## Data Availability

The dataset of the STEPS survey that was used in this research is available at the Ministry of Health upon request to Bicaba Brice: or Zoma Torez ). All survey materials are available on the WHO website (https://extranet.who.int/ncdsmicrodata/index.php/catalog)

## ACKNOWLEDGEMENTS

The authors acknowledge Severin Samadoulougou and Bonkoungou Michel for their contribution to data management.

## AUTHOR CONTRIBUTIONS

KC and MO analyzed data. KC and SS wrote the first draft of the manuscript with inputs of MO, SK, and FKS. FKS and SS conceptualized, formulated research goals, objectives of the study, and the lead methodology. FKS and SK contributed to the acquisition of project financial support. All authors reviewed, edited, and approved the manuscript.

## ETHICS APPROVAL

Before collecting data in the field, the survey protocol was approved by the Ministry of Health’s Ethics Committee for Health Research, and the Ministry of Scientific Research and Innovation, and informed consent was required before the participation of any individual selected for the investigation (**Deliberation No. 2012-12-092 of December, 05, 2012**). The confidentiality of the information collected has been mentioned in the informed consent form.

## CONFLITS OF INTERESTS

Authors declare no conflict of interest.

## REFERENCES

1 World Health Organization. WHO steps surveillance manual: the WHO stepwise approach to chronic disease risk factor surveillance. Geneva: : WHO 2005.

2 Health Effects of Overweight and Obesity in 195 Countries over 25 Years. New England Journal of Medicine 2017;377:13–27. doi:10.1056/NEJMoa1614362

3 Després Jean-Pierre. Body Fat Distribution and Risk of Cardiovascular Disease. Circulation 2012;126:1301–13. doi:10.1161/CIRCULATIONAHA.111.067264

4 Keys A, Fidanza F, Karvonen MJ, et al. Indices of relative weight and obesity. Journal of Chronic Diseases 1972;25:329–43. doi:10.1016/0021-9681(72)90027-6

5 World Health Organization. Waist circumference and waist-hip ratio: report of a WHO expert consultation, Geneva, 8-11 December 2008. Geneva: : World Health Organization 2011.

6 Park J, Lee ES, Lee DY, et al. Waist Circumference as a Marker of Obesity Is More Predictive of Coronary Artery Calcification than Body Mass Index in Apparently Healthy Korean Adults: The Kangbuk Samsung Health Study. Endocrinology and Metabolism 2016;31:559. doi:10.3803/EnM.2016.31.4.559

7 Riera-Fortuny C, Real JT, Chaves FJ, et al. The relation between obesity, abdominal fat deposit and the angiotensin-converting enzyme gene I/D polymorphism and its association with coronary heart disease. Int J Obes (Lond) 2005;29:78–84. doi:10.1038/sj.ijo.0802829

8 Payab M, Amoli MM, Qorbani M, et al. Adiponectin gene variants and abdominal obesity in an Iranian population. Eat Weight Disord 2017;22:85–90. doi:10.1007/s40519-016-0252-1

9 Fu LW, Zhang MX, Wu LJ, et al. [Gene-gene interaction on central obesity in school-aged children in China]. Zhonghua Liu Xing Bing Xue Za Zhi 2017;38:883–8. doi:10.3760/cma.j.issn.0254-6450.2017.07.007

10 Antúnez-Ortiz DL, Flores-Alfaro E, Burguete-García AI, et al. Copy Number Variations in Candidate Genes and Intergenic Regions Affect Body Mass Index and Abdominal Obesity in Mexican Children. BioMed Research International. 2017;2017:e2432957. doi:https://doi.org/10.1155/2017/2432957

11 Zou Y, Ning T, Shi J, et al. Association of a gain-of-function variant in LGR4 with central obesity. Obesity 2017;25:252–60. doi:10.1002/oby.21704

12 Ye D, Cai S, Jiang X, et al. Associations of polymorphisms in circadian genes with abdominal obesity in Chinese adult population. Obesity Research & Clinical Practice 2016;10:S133–41. doi:10.1016/j.orcp.2016.02.002

13 Huang T, Qi Q, Zheng Y, et al. Genetic Predisposition to Central Obesity and Risk of Type 2 Diabetes: Two Independent Cohort Studies. Diabetes Care 2015;38:1306–11. doi:10.2337/dc14-3084

14 Björntorp P, Rosmond R. Neuroendocrine abnormalities in visceral obesity. International Journal of Obesity 2000;24:S80–5. doi:10.1038/sj.ijo.0801285

15 Swinburn BA, Sacks G, Hall KD, et al. The global obesity pandemic: shaped by global drivers and local environments. The Lancet 2011;378:804–14. doi:10.1016/S0140-6736(11)60813-1

16 Celis-Morales C, Livingstone KM, Affleck A, et al. Correlates of overall and central obesity in adults from seven European countries: findings from the Food4Me Study. European Journal of Clinical Nutrition 2018;72:207–19. doi:10.1038/s41430-017-0004-y

17 Sikorski C, Luppa M, Weyerer S, et al. Obesity and Associated Lifestyle in a Large Sample of Multi-Morbid German Primary Care Attendees. PLOS ONE 2014;9:e102587. doi:10.1371/journal.pone.0102587

18 Ross R, Neeland IJ, Yamashita S, et al. Waist circumference as a vital sign in clinical practice: a Consensus Statement from the IAS and ICCR Working Group on Visceral Obesity. Nature Reviews Endocrinology 2020;16:177–89. doi:10.1038/s41574-019-0310-7

19 Zhang Cuilin, Rexrode Kathryn M., van Dam Rob M., et al. Abdominal Obesity and the Risk of All-Cause, Cardiovascular, and Cancer Mortality. Circulation 2008;117:1658–67. doi:10.1161/CIRCULATIONAHA.107.739714

20 Chang J-W, Chen H-L, Su H-J, et al. Abdominal Obesity and Insulin Resistance in People Exposed to Moderate-to-High Levels of Dioxin. PLoS One 2016;11. doi:10.1371/journal.pone.0145818

21 Nam GE, Cho KH, Han K, et al. Obesity, abdominal obesity and subsequent risk of kidney cancer: a cohort study of 23.3 million East Asians. Br J Cancer 2019;121:271–7. doi:10.1038/s41416-019-0500-z

22 Alberti G, Zimmet P, Shaw J, et al. The IDF consensus worldwide definition of the Metabolic Syndrome. 2006.https://www.idf.org/e-library/consensus-statements/60-idfconsensus-worldwide-definitionof-the-metabolic-syndrome.html (accessed 25 Mar 2020).

23 Ezenwaka CE, Okoye O, Esonwune C, et al. High prevalence of abdominal obesity increases the risk of the metabolic syndrome in Nigerian type 2 diabetes patients: using the International Diabetes Federation worldwide definition. Metab Syndr Relat Disord 2014;12:277–82. doi:10.1089/met.2013.0139

24 Després J-P, Lemieux I. Abdominal obesity and metabolic syndrome. Nature 2006;444:881–7. doi:10.1038/nature05488

25 Després J-P. Excess Visceral Adipose Tissue/Ectopic Fat: The Missing Link in the Obesity Paradox?□□Editorials published in the Journal of the American College of Cardiology reflect the views of the authors and do not necessarily represent the views of JACC or the American College of Cardiology. Journal of the American College of Cardiology 2011;57:1887–9. doi:10.1016/j.jacc.2010.10.063

26 Huxley R, Mendis S, Zheleznyakov E, et al. Body mass index, waist circumference and waist:hip ratio as predictors of cardiovascular risk—a review of the literature. European Journal of Clinical Nutrition 2010;64:16–22. doi:10.1038/ejcn.2009.68

27 World Health Organization. Waist circumference and waist-hip ratio: report of a WHO expert consultation, Geneva, 8-11 December 2008. Geneva: : World Health Organization 2011.

28 Wong MCS, Huang J, Wang J, et al. Global, regional and time-trend prevalence of central obesity: a systematic review and meta-analysis of 13.2 million subjects. Eur J Epidemiol 2020;35:673–83. doi:10.1007/s10654-020-00650-3

29 Owolabi EO, Ter Goon D, Adeniyi OV. Central obesity and normal-weight central obesity among adults attending healthcare facilities in Buffalo City Metropolitan Municipality, South Africa: a cross-sectional study. J Health Popul Nutr 2017;36. doi:10.1186/s41043-017-0133-x

30 Mohamed SF, Haregu TN, Khayeka-Wandabwa C, et al. Magnitude and predictors of normal-weight central obesity– the AWI-Gen study findings. Global Health Action 2019;12:1685809. doi:10.1080/16549716.2019.1685809

31 Kabwama SN, Kirunda B, Mutungi G, et al. Prevalence and correlates of abdominal obesity among adults in Uganda: findings from a national cross-sectional, population based survey 2014. BMC Obes 2018;5. doi:10.1186/s40608-018-0217-1

32 Olatunbosun ST, Kaufman JS, Bella AF. Central Obesity in Africans: Anthropometric Assessment of Abdominal Adiposity and its Predictors in Urban Nigerians. Journal of the National Medical Association 2018;110:519–27. doi:10.1016/j.jnma.2018.01.001

33 K. Malik S, Kouame J, Gbane M, et al. Prevalence of abdominal obesity and its correlates among adults in a peri-urban population of West Africa. AIMS Public Health 2019;6:334–44. doi:10.3934/publichealth.2019.3.334

34 Kaboré S, Millogo T, Soubeiga JK, et al. Prevalence and risk factors for overweight and obesity: a cross-sectional countrywide study in Burkina Faso. BMJ Open 2020;10:e032953. doi:10.1136/bmjopen-2019-032953

35 Zeba AN, Delisle HF, Renier G. Dietary patterns and physical inactivity, two contributing factors to the double burden of malnutrition among adults in Burkina Faso, West Africa. Journal of Nutritional Science 2014;3. doi:10.1017/jns.2014.11

36 Marceline YT, Issiaka S, Gilberte KC, et al. Diagnostic et prévalence du syndrome métabolique chez les diabétiques suivis dans un contexte de ressources limitées: cas du Burkina-Faso. Pan Afr Med J 2014;19. doi:10.11604/pamj.2014.19.364.3741

37 Zeba AN, Delisle HF, Renier G, et al. The double burden of malnutrition and cardiometabolic risk widens the gender and socio-economic health gap: a study among adults in Burkina Faso (West Africa). Public Health Nutrition 2012;15:2210–9. doi:10.1017/S1368980012000729

38 Ministère de la santé. Rapport de l’enquête nationale sur la prevalence des principauc facteurs de risques communs aux maladies non transmissibles au Burkina Faso. 2014. https://www.who.int/ncds/surveillance/steps/BurkinaFaso_2013_STEPS_Report.pdf

39 Lean ME, Han TS, Morrison CE. Waist circumference as a measure for indicating need for weight management. BMJ 1995;311:158–61.

40 Tabassum F, Batty GD. Are Current UK National Institute for Health and Clinical Excellence (NICE) Obesity Risk Guidelines Useful? Cross-Sectional Associations with Cardiovascular Disease Risk Factors in a Large, Representative English Population. PLOS ONE 2013;8:e67764. doi:10.1371/journal.pone.0067764

41 Otang-Mbeng W, Otunola GA, Afolayan AJ. Lifestyle factors and co-morbidities associated with obesity and overweight in Nkonkobe Municipality of the Eastern Cape, South Africa. Journal of Health, Population and Nutrition 2017;36:22. doi:10.1186/s41043-017-0098-9

42 INSD. Projection démographique de 2007 à 2020 par région et province. 2009.

43 Yayehd K, Pessinaba S, N’cho-Mottoh MPB, et al. Prevalence and Determinants of Adult Obesity in a Low-Income Population of Western Africa: Results from a Nationwide Cross-Sectional Survey. Obesity & Control Therapies: Open Access 2017;4.https://symbiosisonlinepublishing.com/obesity-control-therapies/obesity-control-therapies33.php (accessed 23 Nov 2020).

44 Agyemang C, Meeks K, Beune E, et al. Obesity and type 2 diabetes in sub-Saharan Africans – Is the burden in today’s Africa similar to African migrants in Europe? The RODAM study. BMC Med 2016;14:1–12. doi:10.1186/s12916-016-0709-0

45 Bender R. Effect of Age on Excess Mortality in Obesity. JAMA 1999;281:1498. doi:10.1001/jama.281.16.1498

46 Sarlio-Lähteenkorva S, Silventoinen K, Lahti-Koski M, et al. Socio-economic status and abdominal obesity among Finnish adults from 1992 to 2002. International Journal of Obesity 2006;30:1653– 60. doi:10.1038/sj.ijo.0803319

47 Yoon YS, Oh SW, Park HS. Socioeconomic status in relation to obesity and abdominal obesity in Korean adults: a focus on sex differences. Obesity (Silver Spring) 2006;14:909–19. doi:10.1038/oby.2006.105

48 Sun Y, Liu B, Snetselaar LG, et al. Association of Normal-Weight Central Obesity With All-Cause and Cause-Specific Mortality Among Postmenopausal Women. JAMA Network Open 2019;2:e197337. doi:10.1001/jamanetworkopen.2019.7337

49 Pischon T, Boeing H, Hoffmann K, et al. General and Abdominal Adiposity and Risk of Death in Europe. New England Journal of Medicine 2008;359:2105–20. doi:10.1056/NEJMoa0801891

50 Fezeu L, Balkau B, Kengne A-P, et al. Metabolic syndrome in a sub-Saharan African setting: central obesity may be the key determinant. Atherosclerosis 2007;193:70–6. doi:10.1016/j.atherosclerosis.2006.08.037

